# Variants in *DNAJC13* Are Not Associated with Parkinson’s Disease Across Different Ancestral Backgrounds

**DOI:** 10.64898/2025.12.30.25343211

**Authors:** César Luis Ávila, Mariam Isayan, Yasser Mecheri, Paula Saffie-Awad, María Milagros Leila, Laurel A Screven, Hampton Leonard, Maria Teresa Periñan, Kristin Levine, Mary B Makarious, the Global Parkinson’s Genetics Program (GP2)

**Author notes:** **Correspondence to** Mary B Makarious, DataTecnica, LLC, Washington DC, USA 20037, **E-mail:**. Funding sources for study Data used in the preparation of this article were obtained from Global Parkinson’s Genetics Program (GP2; https://gp2.org). GP2 is funded by the Aligning Science Across Parkinson’s (ASAP) initiative and implemented by The Michael J. Fox Foundation for Parkinson’s Research. Additional funding was provided by The Michael J. Fox Foundation for Parkinson’s Research through grant MJFF-009421/17483. The AMP® PD program is a public-private partnership managed by the Foundation for the National Institutes of Health and funded by the National Institute of Neurological Disorders and Stroke (NINDS) in partnership with the Aligning Science Across Parkinson’s (ASAP) initiative; Celgene Corporation, a subsidiary of Bristol-Myers Squibb Company; GlaxoSmithKline plc (GSK); The Michael J. Fox Foundation for Parkinson’s Research; Pfizer Inc.; Sanofi US Services Inc.; and Verily Life Sciences.

## Abstract

**Background:** *DNAJC13* was initially linked to autosomal dominant (AD) Parkinson’s disease (PD) in a European Mennonite family carrying the p.N855S variant. However, imperfect segregation and conflicting reports of pathogenicity raised uncertainty of *DNAJC13*’s role in the disease.

**Objectives:** Explore the association between common and rare variants in *DNAJC13* and Parkinson’s disease.

**Methods:** We leveraged the largest available PD genetics data from the Accelerating Medicines Partnership - Parkinson Disease (AMP-PD) and the diverse ancestry available through the Global Parkinson’s Genetics Program (GP2), consisting of 2,471 patients and 3,098 controls, and 44,186 patients and 27,066 controls, respectively, to perform burden tests and association tests for rare and common variants, respectively.

**Results:** Burden analysis showed no association between rare variants in *DNAJC13* and PD. However, association analysis within common non-synonymous variants nominated 5 variants within *DNAJC13*. Nevertheless, these associations require further validation since the analyses are still underpowered.

**Discussion:** Our analysis did not find further evidence supporting *DNAJC13* involvement in PD. However, studies of even larger cohorts and AD-PD families may bring definite answers about *DNAJC13’s* role in PD.

## Introduction

Vilariño-Güell and colleagues (1), linked a missense variant in the *DNAJC13* gene (p.N855S, rs387907571) to autosomal dominant late-onset Parkinson’s disease (PD) in a large Dutch-German-Russian Mennonite family.

The *DNAJC13* gene or RME-8 (receptor-mediated endocytosis 8) encodes a heat shock protein with a co-chaperone domain that has been found to regulate clathrin dynamics on early endosomes (2). More recently, it was suggested that *DNAJC13* is connected to Autophagic Lysosome Reformation (ALR) and autophagy and could play a role in neuronal health and the progression of neurodegenerative diseases (3).

In the subsequent years, several smaller families and individual patients with *DNAJC13* variants have been documented (4–8) and some functional studies have supported a gene-disease relationship (9–11). In addition, other DNAJC proteins have been suggested to increase the risk of developing PD, such as DNAJC26 and DNAJC10 (12). Consequently, the evidence for *DNAJC13*’s role in PD remains under debate.

We leverage the large cohort of patients with PD and controls recruited through the Accelerating Medicines Partnership - Parkinson Disease (AMP-PD) and the Global Parkinson’s Genetics Program (GP2) (Global Parkinson’s Genetics Program 2021) to study the association between non-synonymous coding variants in *DNAJC13* and PD.

## Methods

To investigate the association between *DNAJC13* and PD, we first analyzed whole-genome sequencing (WGS) data obtained from the AMP-PD initiative (https://amp-pd.org/) release 4.0, consisting of 2,471 patients and 3,098 controls from European and Ashkenazi Jewish descent (**Supplementary Table 1**). Additionally, we used large-scale genotyping imputed data from GP2, consisting of 44,186 PD patients and 27,066 controls from nine different ancestry populations: European [EUR], African Admixed [AAC], African [AFR], Ashkenazi Jews [AJ], Admixed American/Latin American [AMR], Central Asian [CAS], East Asian [EAS], Middle Eastern [MDE], South Asian [SAS]. Quality control analyses at a sample and variant level are described elsewhere [https://github.com/GP2code/GenoTools] (13).

Common variants (MAF > 0.01) were annotated using ANNOVAR (14), focusing on non-synonymous variants. Fisher’s exact test was applied to examine the differences between allele frequencies using PLINK 1.9 (15). SNP-phenotype association analyses were performed with a generalized linear model and adjusted for multiple comparisons using Bonferroni correction method as implemented in the PLINK 2.0 software package (16,17). The number of variants included in Bonferroni correction corresponds to the number of non-synonymous exonic variants found within each ancestry (**Supplementary Table 2**). In the logistic regression analysis, we included sex and the first five genetic principal components (PCs) as covariates to account for population stratification. We used the LDlink tool (18) to assess linkage disequilibrium between potential relevant variants identified in our analysis and the previously identified SNPs in chromosome 3 according to the largest genome-wide meta-analysis in European population performed by Nalls and colleagues (19) and the multi-ancestry meta-analysis by Kim et al. (20). Finally, to assess the cumulative effect of multiple low frequency variants (MAF < 0.01) on the risk for PD, gene-based burden analyses were performed using RVTESTS (21).

## Data Sharing

Data used in the preparation of this article were obtained from GP2. Specifically, we used Tier 2 data from GP2 (release 11; DOI 10.5281/zenodo.17753486). GP2 data can be accessed through AMP PD (https://amp-pd.org). Genotyping imputation, quality control, ancestry prediction, and processing was performed using GenoTools v1.0, publicly available on GitHub (https://github.com/GP2code/GenoTools). All scripts for analyses are publicly available on GitHub (https://github.com/GP2code/DNAJC13-PD-GeneBurden; DOI: 10.5281/zenodo.18063253).

All cohorts recruited to GP2 undergo a thorough review of the consent forms, ensuring that each contributing study abided by the ethics guidelines set out by their institutional review boards, and all participants gave informed consent for inclusion in both their initial cohorts and subsequent studies within local law constraints. All GP2 data is hosted in collaboration with the Accelerating Medicines Partnership in Parkinson’s disease, and is available via application on the website (https://amp-pd.org/register-for-amp-pd).

## Results

We identified a total of 501 variants (MAF >1%) within the *DNAJC13* gene in the European AMP-PD WGS data, of which 8 were exonic, including 3 synonymous and 5 missense variants (**Supplementary Table 2**). Two variants in this dataset were associated with PD risk, i.e. p.A1421V (rs61748102) and p.1463S (rs3762672). However, only p.A1463S was still significant after Bonferroni correction. This variant was enriched in healthy controls compared to PD patients (Beta = -0.102, SE = 0.040; p-value BONF = 0.01) (**Table 1**). We were unable to find any significant association between common variants in DNAJC13 and PD in the AJ ancestry.

**Table 1:**
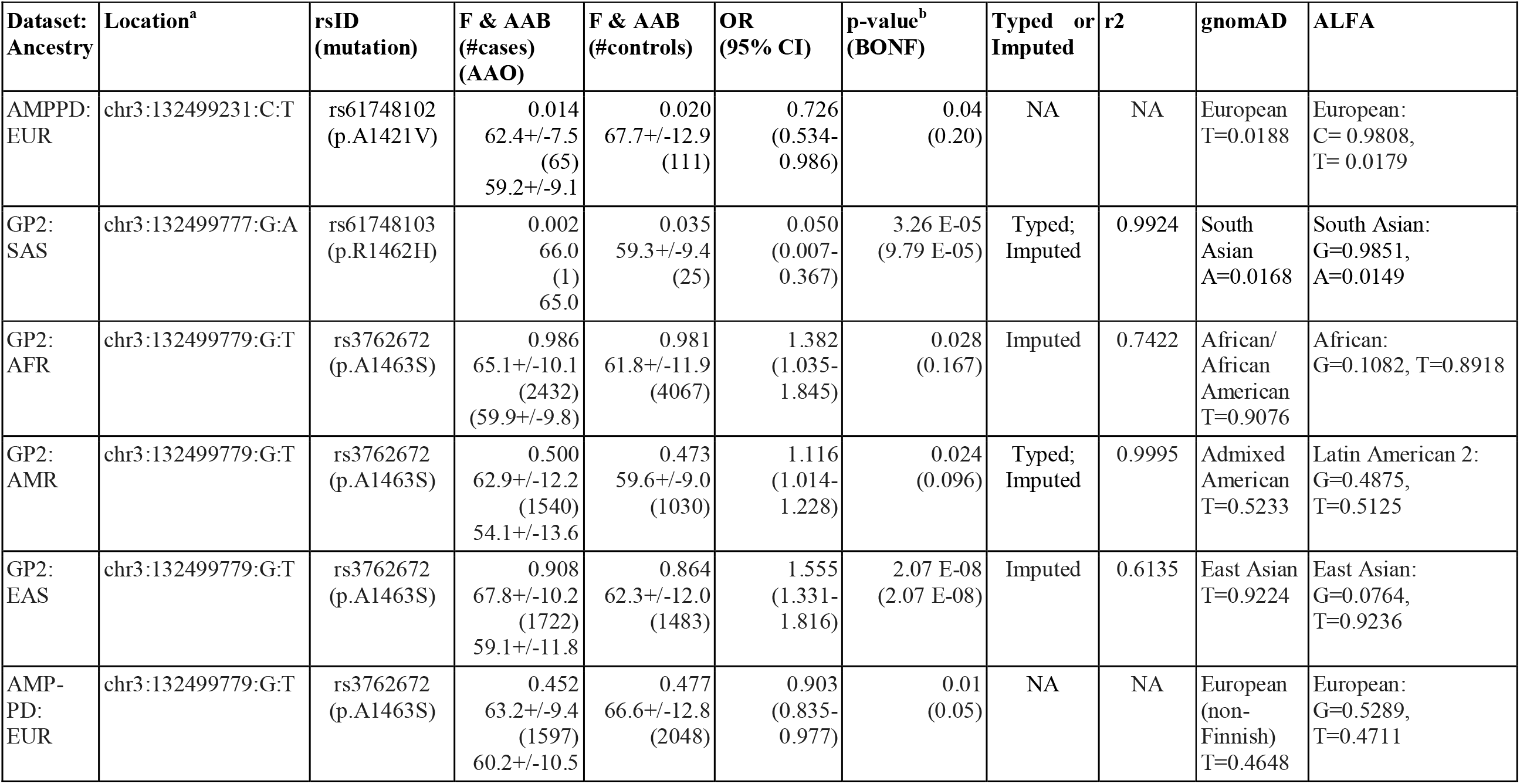
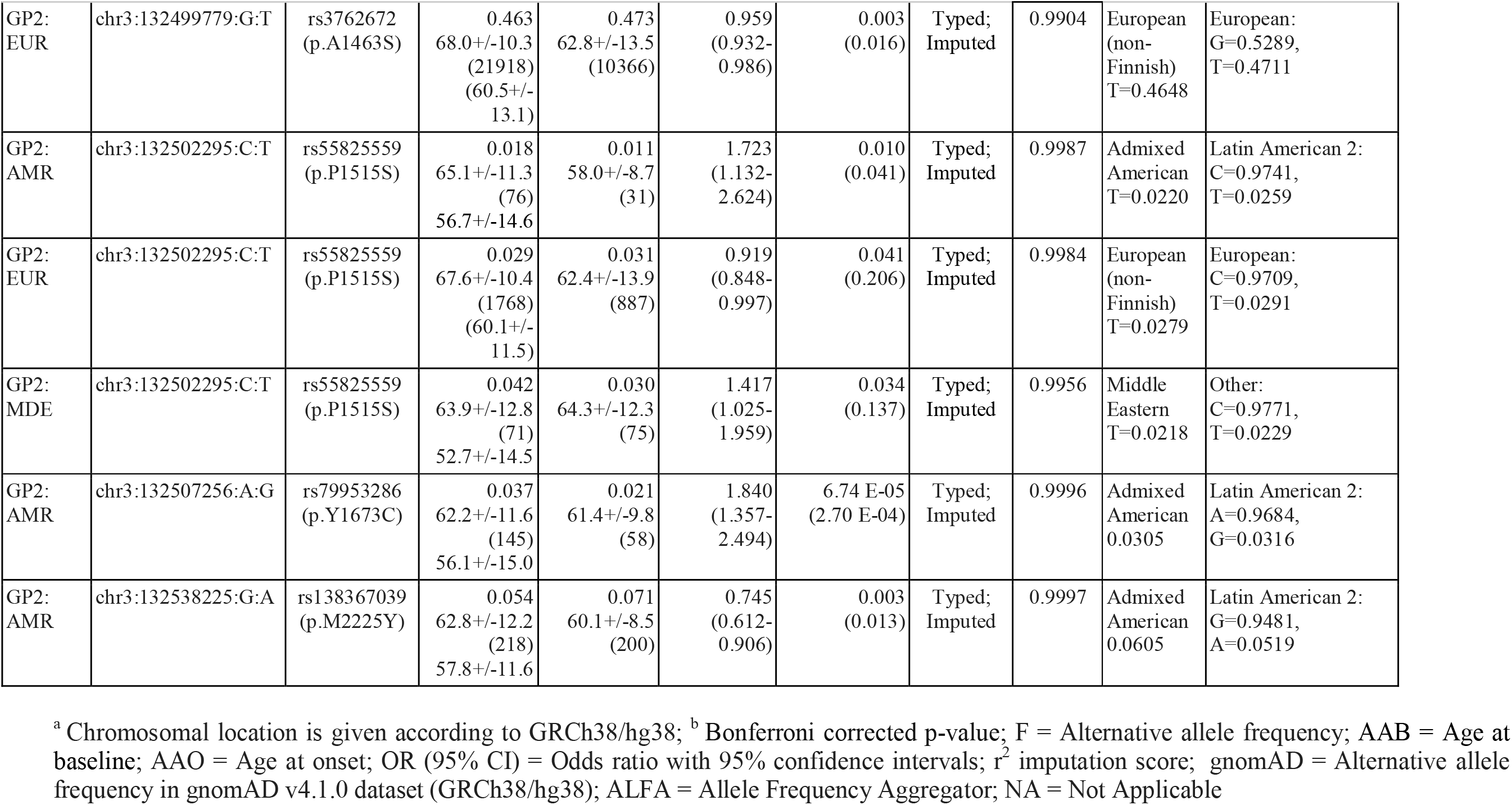
Non-synonymous exonic variants observed in *DNAJC13* (NM_001329126.2) based on case-control frequencies across populations, selected by significance threshold of p<0.05.

In the GP2 genotyping dataset we evaluated a total number of 10 unique exonic non-synonymous common variants across the different ancestries. Among them, the p.A1463S (rs3762672) variant was found once again enriched in healthy controls in the EUR ancestry (Beta = -0.042; SE =0.014; p-value = 0.003). On the contrary, the same mutation was enriched in PD patients within the AFR (Beta = 0.324; SE=0.147; p-value = 0.028), EAS (Beta = 0.441; SE=0.079; p-value = 2.07 E-08) and AMR (Beta = 0.110; SE=0.049; p-value = 0.024) ancestries, but only EAS remained significant after Bonferroni correction. In addition to p.A1463S, other four missense variants exhibited significant p-values after Bonferroni correction (**Table 1**): p.R1462H (rs61748103) in SAS (Beta = -2.996; SE = 1.010; p-value = 3.26 E-05) and p.P1515S (rs55825559) (Beta = 0.544; SE = 0.214; p-value = 0.010), p.Y1673C (rs79953286) (Beta = 0.610; SE = 0.155; p-value= 6.74 E-05) and p.M2225Y (rs138367039) (Beta = - 0.294; SE = 0.100 ; p-value= 0.003) in AMR. None of these variants were in linkage disequilibrium with SNPs identified in chromosome 3 according to the genome-wide meta-analyses performed by Nalls et al. (19) and Kim et al. (20) (rs73038319, rs6808178, rs10513789, rs55961674, rs11707416, rs1450522, and rs10513789). Still, these results should be taken with caution since they might be underpowered.

We were unable to evaluate the association between PD and individual variants reported in familial cases such as p.N855S (rs387907571, MAF=2.03 E-05), p.T1895M (rs145242123; MAF = 3.09 E-03), or p.V722L (rs146930051, MAF = 7.25 E-05), due to their very low representation within the GP2 dataset. Also, the p.P336A, and p.R1266Q variants with unknown pathogenicity identified through sequencing in the Canadian population by Gustavsson and colleagues (22) were absent in the GP2 dataset. Nevertheless, burden analysis showed no association between rare variants and PD (**Supplementary Table 3**).

## Discussion

The aim of our study was to investigate whether genetic variation within *DNAJC13* is associated with increased PD risk by analyzing large genetic datasets from the AMP-PD and GP2 initiatives. Similar to previous studies performed in Caucasians (n=1938 PD cases) (23), French-Canadian/French (n=528 PD cases) (24) and Italians (5) (n=731 PD cases) our analysis did not find a significant association between rare variants of *DNAJC13* and PD. This can be explained by their ultra rare frequency across the different ancestries. For instance, the p.N855S variant was only reported in five PD patients (1,22), four of whom were from the original Mennonite family. Additionally, the pathogenicity of the p.N855S variant remains questionable, as it had an incomplete segregation with the disease within the index family and an independent reanalysis identified the *TMEM230* p.A141L variant to be the only fully co-segregating mutation in the entire family (25). Of the four rare missense variants (p.P336A, p.V722L, p.R1266Q, and p.T1895M) with unknown pathogenicity previously reported by Gustavsson et. al (22) to only be present in PD cases in a Canadian-Norwegian cohort, only p.T1895M and p.V722L were present in our dataset. Our findings on the aggregated impact of rare variants on PD is in accordance with previous reports based on the analysis of EUR and AJ ancestries within the McGill, Columbia, and Sheba cohorts (26).

Leveraging AMP-PD and GP2’s dataset that includes diverse populations allowed us to identify six common variants associated with PD (p<0.05), however only five remained significant after multiple test correction. The p.A1463S variant (rs3762672) is enriched in the AMP-PD and GP2 dataset in healthy controls in the EUR ancestry but its impact on PD risk seems to be very mild (OR=0.90 and OR=0.95, respectively). The significance of this variant becomes even more puzzling in the light of the findings within the AMR, AFR and EAS ancestries, where the mutation is enriched in patients vs controls. Accordingly, this variant is reported in ClinVar as benign based on two reports (RCV001579441.2, RCV001619990.3).

The p.R1462H (rs61748103) and p.M2225Y (rs138367039) were found to be enriched in controls within the SAS (F_affected_=0.002, F_unaffected_=0.033, p-value=2.67 E-05) and AMR ancestries (F_affected_=0.054, F_unaffected_=0.072, p-value=0.003), respectively. The p.R1462H variant was not reported in ClinVar at the time of writing. The p.P1515S and p.Y1673C variants were enriched in PD patients within the AMR ancestry. However, the sample size in these ancestries was very small, and we could not replicate these findings in other ancestries. Further functional studies should be performed to assess the pathological impact of these mutations.

Overall, after analyzing both GP2 multi ancestry genotyping and AMP-PD WGS datasets, we were unable to identify any significant association between PD risk and variants in *DNAJC13* or to replicate previously reported associations. However, focused larger ancestry specific studies and family studies may be useful to identify ultra-rare variants segregating with the disease.

## Data Availability

Data used in the preparation of this article were obtained from GP2. Specifically, we used Tier 2 data from GP2 (release 11; DOI 10.5281/zenodo.17753486). GP2 data can be accessed through AMP PD (https://amp-pd.org). Genotyping imputation, quality control, ancestry prediction, and processing was performed using GenoTools v1.0, publicly available on GitHub (https://github.com/GP2code/GenoTools). All scripts for analyses are publicly available on GitHub (https://github.com/GP2code/DNAJC13-PD-GeneBurden; DOI: 10.5281/zenodo.18063253).
All cohorts recruited to GP2 undergo a thorough review of the consent forms, ensuring that each contributing study abided by the ethics guidelines set out by their institutional review boards, and all participants gave informed consent for inclusion in both their initial cohorts and subsequent studies within local law constraints. All GP2 data is hosted in collaboration with the Accelerating Medicines Partnership in Parkinson's disease, and is available via application on the website (https://amp-pd.org/register-for-amp-pd).

https://amp-pd.org/register-for-amp-pd

## Acknowledgements

This work was carried out with the support and guidance of the “GP2 Trainee Network” which is part of the Global Parkinson’s Genetics Program (GP2; https://gp2.org). GP2 is funded by the Aligning Science Across Parkinson’s (ASAP) initiative and implemented by The Michael J. Fox Foundation for Parkinson’s Research (MJFF). For a complete list of GP2 members see https://doi.org/10.5281/zenodo.7904831.

## Author Roles

(1) Research Project: A. Conception, B. Organization, C. Execution; (2) Statistical Analysis: A. Design, B. Execution, C. Review and Critique; (3) Manuscript Preparation: A. Writing of the First Draft, B. Review and Critique.

C.L.A.: 1B, 1C, 2B, 3A, 3B; Y.M.: 1B, 1C, 2B, 3A, 3B; M.I.: 1B, 1C, 2B, 3A, 3B; P.S.A.: 1B, 1C, 2B, 3A, 3B; M.L.: 1B, 1C, 2B, 3A, 3B; L.S.: 1B; H.L.: 1A, 1B, 3B; S.B-C: 3B; M.T.P.: 3B; K.L.: 1A, 1B, 2C, 3A, 3B; M.B.M.: 1A, 1B, 1C, 2A, 2C, 3A, 3B

