## Supplemental Information for "Variants in *DNAJC13* Are Not Associated with Parkinson’s Disease Across Different Ancestral Backgrounds"

### Supplementary Information

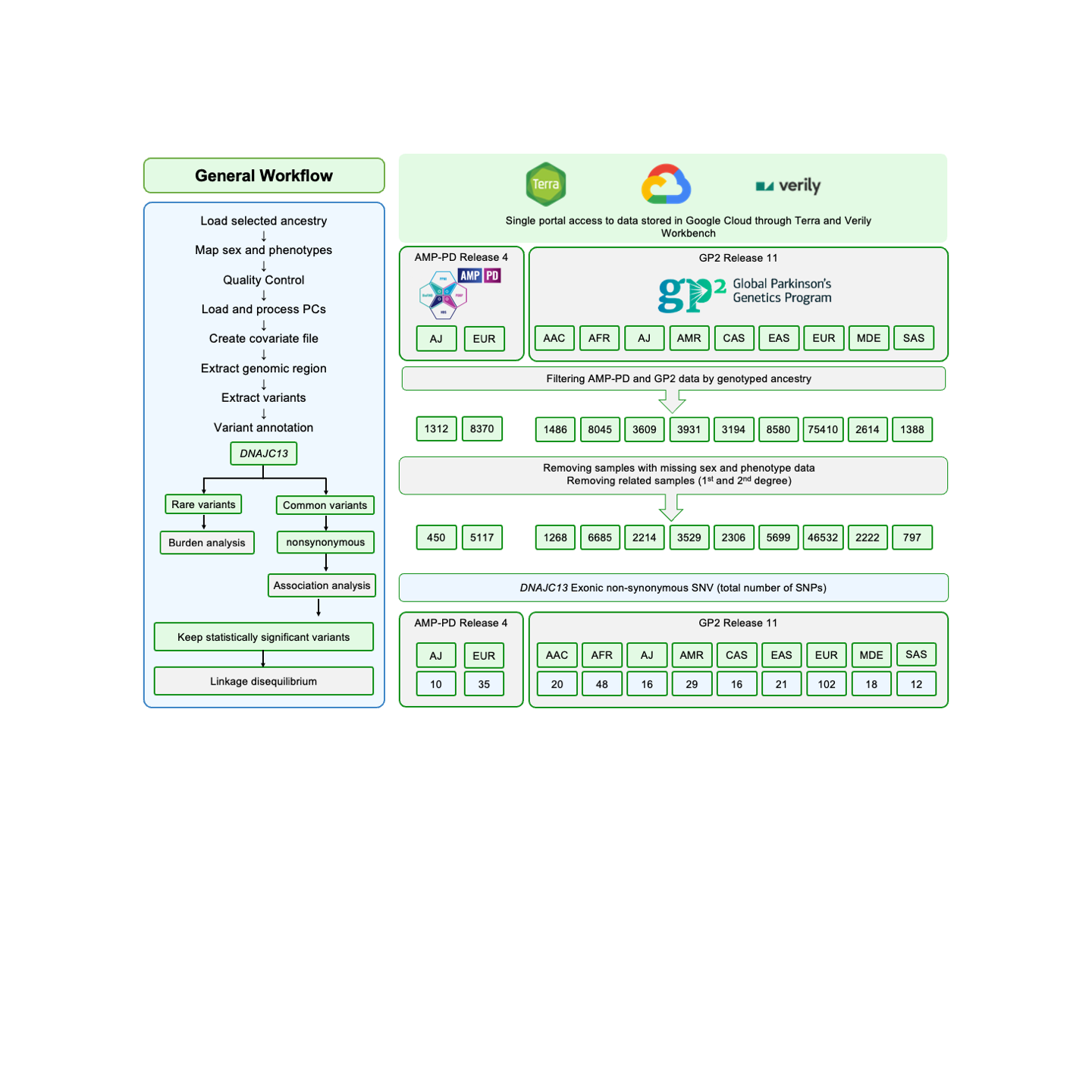

**Figure S1: A comprehensive review of steps undertaken in the *DNAJC13* gene variant extraction and association analysis.**

**Supplementary Table 1: Demographics Information - AMP-PD release 4 and GP2 release 11.**

| Cohort | Ancestry | # of subjects | # Control | | #PD | |
| --- | --- | --- | --- | --- | --- | --- |
|  |  |  | Male | Female | Male | Female |
| AMP-PD | EUR | n_total_= 5117  n_cases_= 2275 n_controls_= 2842 | 1376 | 1466 | 1441 | 834 |
|  | AJ | n_total_=450  n_cases_= 190 n_controls_= 260 | 127 | 133 | 130 | 60 |
| GP2 | AAC | n_total_= 1268  n_cases_= 470 n_controls_= 798 | 286 | 512 | 286 | 184 |
|  | AFR | n_total_= 6685  n_cases_= 2510 n_controls_= 4175 | 2155 | 2020 | 1743 | 767 |
|  | AJ | n_total_= 2214  n_cases_= 1756 n_controls_= 458 | 276 | 182 | 1178 | 578 |
|  | AMR | n_total_= 3529  n_cases_= 2090 n_controls_= 1439 | 504 | 935 | 1215 | 875 |
|  | CAS | n_total_= 2306  n_cases_= 997 n_controls_= 1309 | 535 | 774 | 435 | 562 |
|  | EAS | n_total_= 5699  n_cases_= 3026 n_controls_= 2673 | 1797 | 876 | 1585 | 1441 |
|  | EUR | n_total_= 46532  n_cases_= 31981 n_controls_= 14551 | 7177 | 7374 | 20208 | 11773 |
|  | MDE | n_total_= 2222  n_cases_= 950 n_controls_= 1272 | 561 | 711 | 555 | 395 |
|  | SAS | n_total_= 797  n_cases_= 406 n_controls_= 391 | 273 | 118 | 260 | 146 |

**Supplementary Table 2: *DNAJC13* common variants (MAF > 0.01) identified in the AMP-PD Release 4 and GP2 Release 11 datasets****.**

| **Cohort** | **Ancestry** | **Total variants^a^** | **Intronic** | **3’-UTR** | **5’-UTR** | **Exonic** | |
| --- | --- | --- | --- | --- | --- | --- | --- |
|  |  |  |  |  |  | **Synonymous** | **Nonsynonymous** |
| AMP-PD | EUR | 501 | 491 | 2 | 0 | 3 | 5 |
|  | AJ | 519 | 508 | 3 | 0 | 4 | 4 |
| GP2 | AAC | 506 | 493 | 4 | 1 | 3 | 5 |
|  | AFR | 477 | 464 | 3 | 1 | 3 | 6 |
|  | AJ | 273 | 263 | 3 | 0 | 3 | 4 |
|  | AMR | 249 | 242 | 2 | 0 | 1 | 4 |
|  | CAS | 221 | 214 | 2 | 0 | 1 | 4 |
|  | EAS | 223 | 220 | 2 | 0 | 0 | 1 |
|  | EUR | 256 | 247 | 2 | 0 | 2 | 5 |
|  | MDE | 239 | 232 | 2 | 0 | 1 | 4 |
|  | SAS | 222 | 214 | 4 | 0 | 1 | 3 |

**Supplementary Table 3: Burden analysis on rare variants in GP2 Release 11 dataset**

| **Ancestry** | **Number of rare variants** | **Skat**  **p-Value** | **SkatO**  **p-Value** |
| --- | --- | --- | --- |
| **AAC** | 659 | 0.376 | 0.529 |
| **AFR** | 1352 | 0.155 | 0.269 |
| **AJ** | 430 | 0.895 | 1.000 |
| **AMR** | 1050 | 0.431 | 0.224 |
| **CAS** | 701 | 0.683 | 0.663 |
| **EAS** | 1095 | 0.770 | 0.697 |
| **EUR** | 3827 | 0.183 | 0.303 |
| **MDE** | 735 | 0.225 | 0.361 |
| **SAS** | 489 | 0.384 | 0.545 |
